## Supplemental Table and Figures for "Versatile and flexible microfluidic qPCR test for high-throughput SARS-CoV-2 and cellular response detection in nasopharyngeal swab samples"

**Supplemental Table 1: List of the micro-RNA tested using the Biomark HD system.**

Ctrl_miRTC_1
Hs_let-7a_2
Hs_let-7d_1
Hs_miR-1_2
Hs_miR-103a_1
Hs_miR-10a_2
Hs_miR-122a_1
Hs_miR-125a_1
Hs_miR-125b_1
Hs_miR-126_1
Hs_miR-130a_1
Hs_miR-133a_2
Hs_miR-140_1
Hs_miR-141_1
Hs_miR-142-3p_2
Hs_miR-143_1
Hs_miR-146a_1
Hs_miR-148a_1
Hs_miR-148b_2
Hs_miR-150_1
Hs_miR-152_1
Hs_miR-155_2
Hs_miR-15b_2
Hs_miR-16_2
Hs_miR-17_2
Hs_miR-181a_2
Hs_miR-18a*_1
Hs_miR-191_1
Hs_miR-195_1
Hs_miR-197_2
Hs_miR-199a-3p_1
Hs_miR-200b_3
Hs_miR-200c_1
Hs_miR-203_1
Hs_miR-204_1
Hs_miR-205_1
Hs_miR-20b_1
Hs_miR-21_2
Hs_miR-210_1
Hs_miR-214_2
Hs_miR-216a_1
Hs_miR-22_1
Hs_miR-221_1
Hs_miR-223_1
Hs_miR-23a_2
Hs_miR-23b_2
Hs_miR-24_1
Hs_miR-25_1
Hs_miR-27a_1
Hs_miR-27b_2
Hs_miR-28_1
Hs_miR-296-5p_1
Hs_miR-29a_1
Hs_miR-29b_1
Hs_miR-29c_1
Hs_miR-301a_1
Hs_miR-30b_1
Hs_miR-30d_2
Hs_miR-31_1
Hs_miR-320a_1
Hs_miR-328-3p_1
Hs_miR-331_1
Hs_miR-34a_1
Hs_miR-34b_2
Hs_miR-34c_1
Hs_miR-3620-3p_1
Hs_miR-374a_1
Hs_miR-375_2
Hs_miR-382_2
Hs_miR-409-3p_1
Hs_miR-424_1
Hs_miR-429_1
Hs_miR-4455_1
Hs_miR-449_1
Hs_miR-449b_1
Hs_miR-451_1
Hs_miR-484_1
Hs_miR-486_1
Hs_miR-548b-5p_1
Hs_miR-574-3p_1
Hs_miR-660_1
Hs_miR-92_1
Hs_miR-93_1
Hs_miR-98_1
Hs_RNU6-2_11

### Supplemental Figures

Fassy, Lacoux et al. Fig S1

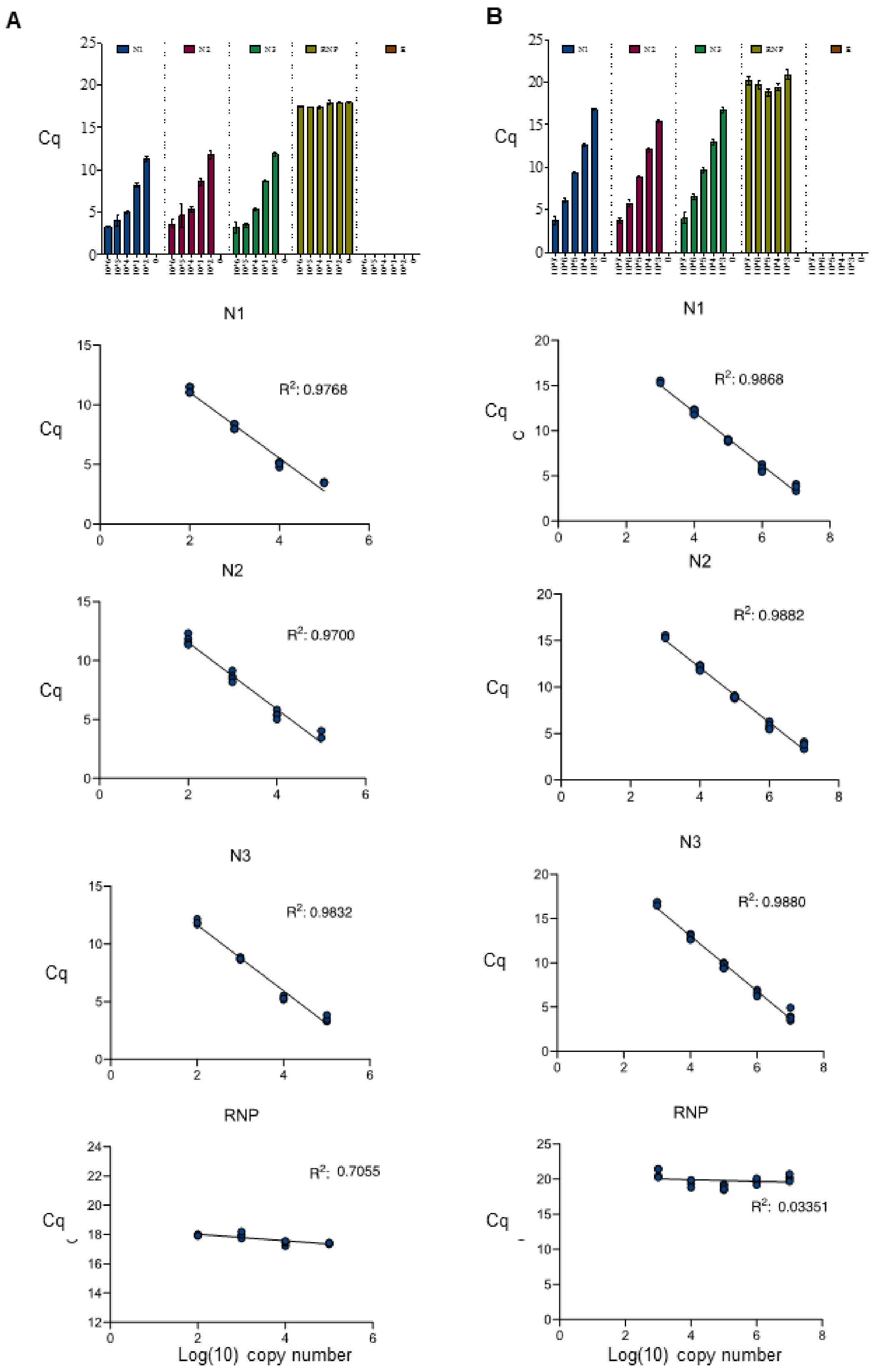

**Fig.S1. Titration of the diluted nucleocapside spike-in transcript.** A ten-fold serial dilution ranging from 1 to $10^{-6}$ was prepared from the stock solution of the in vitro-transcribed N gene and supplemented with 2 ng/µL of total RNA from HEK 293 Cells. Reverse Transcription was performed followed by 15 cycles of pre-Amplification and 30 cycles of qPCR. The RT-qPCR reaction was performed without (**A**) or with (**B**) a RNA purification step**.** Linear regression was performed by logarithmic plots of transcript copy number against Cq value. We observed a good correlation according Cq linear regression curves according to dilution for the three viral CDC primers/probe sets (N1, N2, N3). No Cq value has been detected for the E primers/probe. RNP, used as internal control, shows constant detection of Cq value, suggesting a good performance of the qPCR.

Fassy, Lacoux et al. Fig S2

Correlation study – N probe

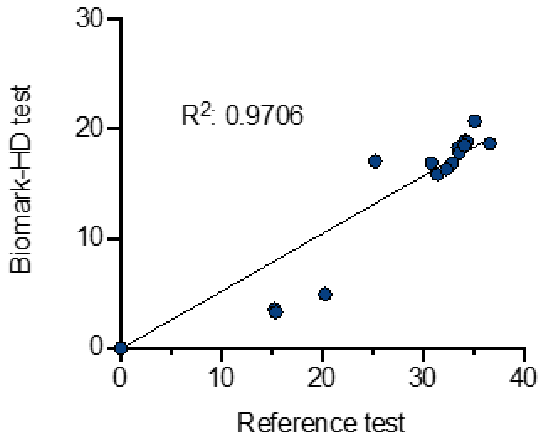


**Fig.S2**. **Validation of the Biomark-HD protocol on a cohort of 92 biopsies including 15 positive patients.** The correlation of the Cq values obtained for the N primers/probe (Biomark-HD) and the GeneFirst COVID-19 detection kit is presented.

Fassy, Lacoux et al. Fig S3

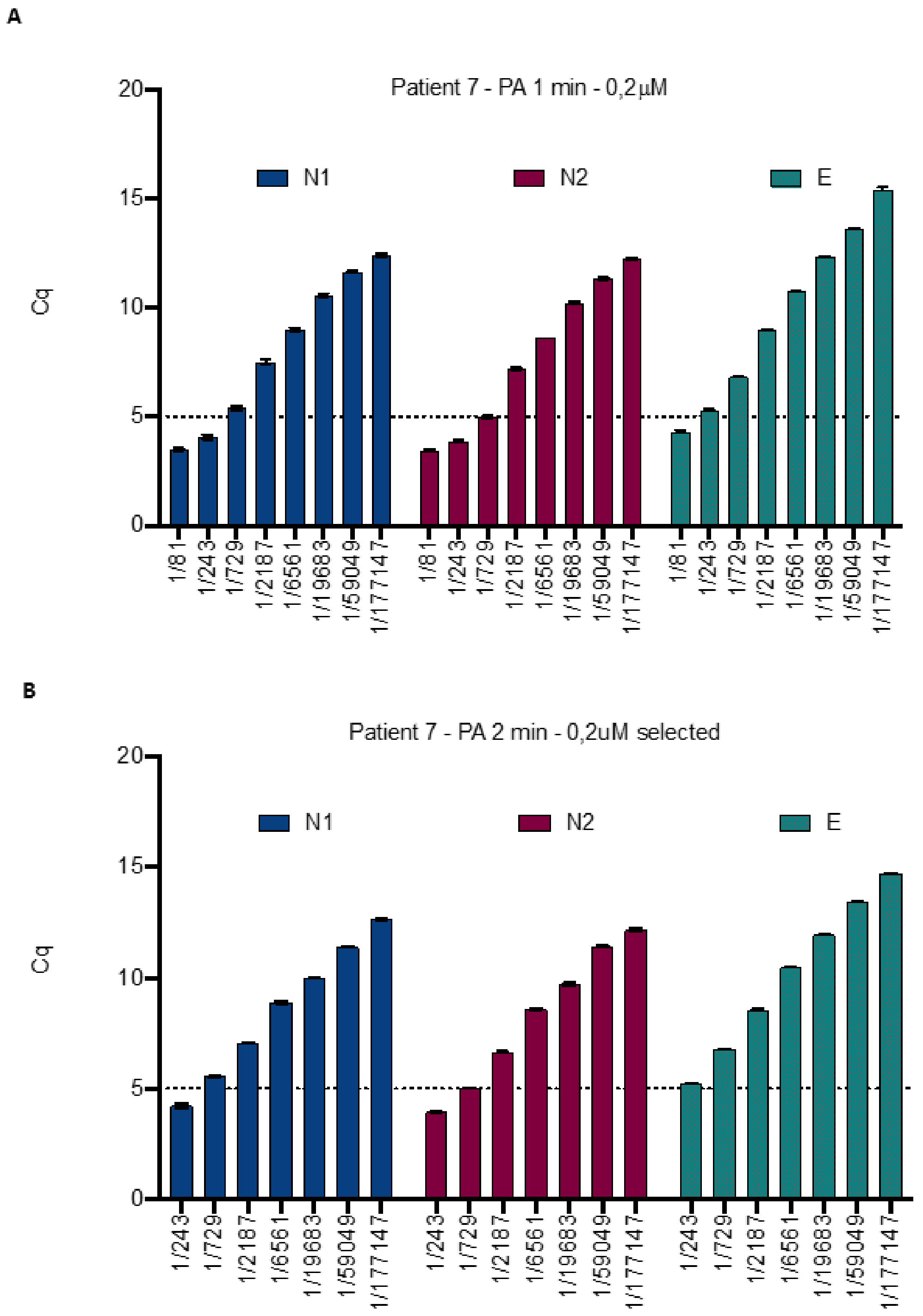


---

**Fig.S3: Preamplification step optimisation**. The elongation time used in the preamplification reaction was reduced to 1 min at 60°C (**A**) from 2 min at 60°C (**B**) using diluted total RNA from a SARS-nCov2 positive patient sample.

Fassy, Lacoux et al. Fig S4

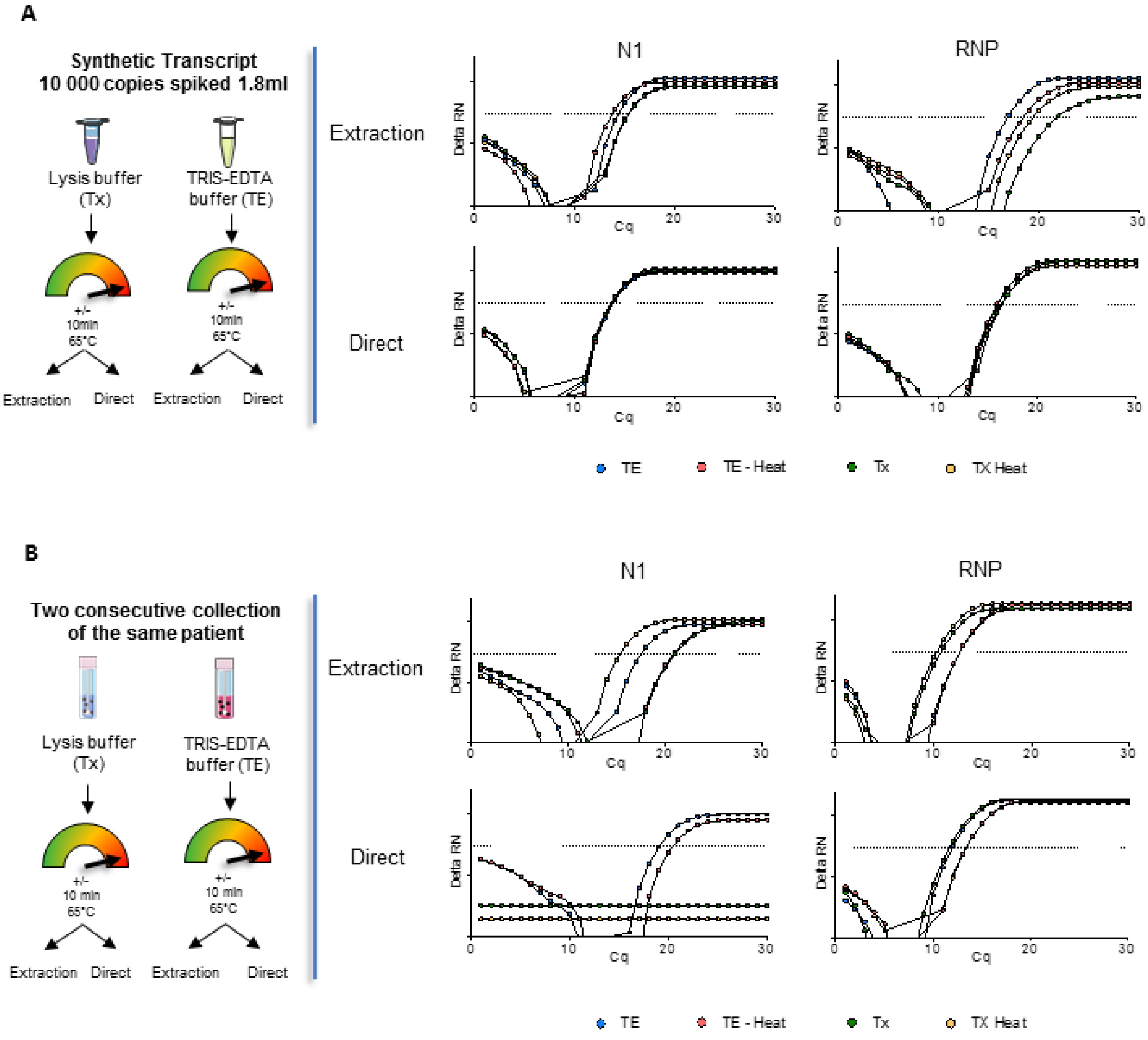


**Fig. S4 Effect of Triton X-100 on extraction-based and direct RT-qPCR protocol performed on a synthetic N transcript or a clinical sample. A.** *In vitro*-transcribed viral N gene was added to either a Triton X-100 containing lysis buffer (Tx) or to TE buffer (TE). The samples were heated or not at 65°C for 10 min. RNA extraction was performed or not (direct) and N1 or RNP levels were determined by RT-qPCR using the Biomark-HD system. **B.** A similar protocol as in A was used but the starting material were two consecutive sample collection from the same patient processed either in a Triton X-100 containing lysis buffer or to TE buffer.

Fassy, Lacoux et al. Fig S5

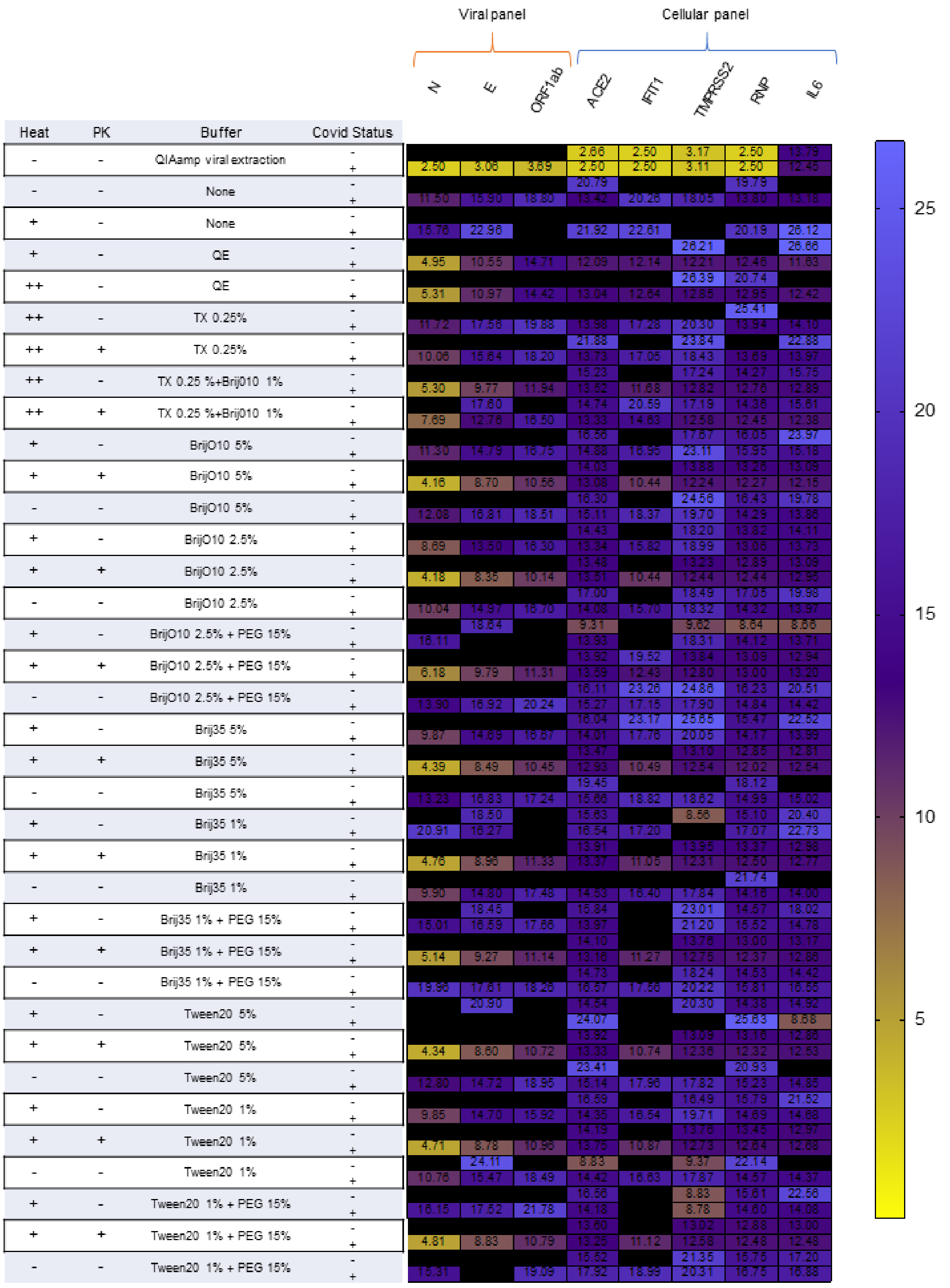

**Fig.S5: Fig.S5: Heatmap of Cq values from direct RT-qPCR experiments using various combinations of detergent on clinical samples.** Samples from a positive- or negative-COVID-19 patient collected in a commercial VTM was mixed with different combinations of detergents/emulsifiers, in presence or absence of PK (2 mg/mL) and further heat at 95°C for 5 min or not. Cq values obtained in quadruplicate are presented. Tx: Triton X100; PEG: poly ethylene glycol 600; QE: Quick Extract™ DNA Extraction Solution.
